# Personalizing supportive healthcare for patients with immunological disorders

**DOI:** 10.1101/2024.04.25.24306403

**Authors:** Tessa S. Folkertsma, Reinhard Bos, Robert M. Vodegel, Sjaak Bloem, Aad R. Liefveld, Greetje J. Tack

## Abstract

Current insights to personalize supportive care for patients with immunological disorders, especially in the context of medical treatments, remain inadequate. Delivering and guiding supportive care unquestionably contributes to a higher quality of life and better overall healthcare. The ‘Subjective Health Experience (SHE) Model’ provides a general framework, comprising four segments, to differentiate supportive healthcare in a quick and practical approach. In this report both health care workers and patients tailored the unique needs of patients with immunological disorders to improve their supportive care.

Employing qualitative methods, group discussions and individual interviews were conducted with 19 healthcare professionals and 18 patients suffering from Rheumatoid Arthritis/Spondylarthritis, Inflammatory Bowel Disease (Crohn’s disease and Ulcerative colitis), and Psoriasis/Hidradenitis Suppurativa. The aim was to ascertain nuanced insights into the behaviour, questions, and needs of patients with six common immunological conditions guided by the SHE-model, thereby refining the personalized supportive care framework.

A detailed description was made for patients with immunological disorders per SHE-model segment. Based on these insights, it was determined for each segment **‘WHAT’** kind of supportive care is needed and **‘HOW’** it should be offered. Notably, patients emphasized the qualitative aspects of their interactions with healthcare professionals (attention, acknowledgment, and empathetic communication), contrasting with professionals’ focus on the treatment plan. This led to a strategic allocation of supportive care interventions across patient segments.

This study has significantly advanced our understanding of appropriate supportive healthcare for patients with immunological disorders from the perspective of the SHE-model. These findings not only enrich the existing literature but also equip healthcare professionals with a concrete guide for enhancement of supportive care, as the SHE-model is easy to perform in daily clinical care. Attention, acknowledgment, and listening comprise the foundational elements for offering and guiding supportive care.

## Introduction

The quality of life (QoL) of individuals with immunological disorders is not only significantly impacted by physical and biomedical complications, but also by the psychological and psychosocial challenges of these conditions. Disease specific complications include joint deformities, uveitis [1], cardiovascular problems, and osteoporosis for Rheumatoid Arthritis (RA) and Spondylarthritis (SpA) [2], malnutrition [3,4], colorectal cancer [5,6], stenosis [7], and fistulas [8] for Inflammatory Bowel Disease (IBD; Cohn’s disease and Ulcerative colitis) and, arthritis [9], cardiovascular diseases [10], and secondary bacterial infections for Psoriasis (PsO) and Hidradenitis Suppurativa (HS) [11]. Psychological issues are often overlapping for the disorders, such as depression, anxiety, stigmatization and social withdrawal [2,11,12]

In addition to appropriate medical treatment, it is essential to consider how individuals experience their own health [13]. Incorporating these experiences with biomedical perspectives can lead to numerous advantages. It can streamline diagnosis, refine treatment plans, enhance patient adherence to therapeutic regimens, and improve patient-reported satisfaction levels [14]. Furthermore, insights into health perception can be used to tailor supportive strategies, ensuring they resonate with the unique needs of each patient [15].

Contemporary healthcare paradigms are progressively gravitating towards the concept of ‘tailored care’ – the right supportive care, at the right time, in the right place. This shift can be attributed to several developments, including the higher healthcare demand due to increased prevalence of chronic (immunological) conditions [16–18]. In addition, the demand for healthcare is becoming more challenging to meet due to healthcare personnel shortages [19]. Within this context and in light of the aforementioned impact on QoL, it is crucial for the treatment of immunological disorders to integrate tailored care, to deploy resources efficiently, and actively engage patients in their care processes, capitalizing on their proactive contributions.

Bloem and Stalpers conceptualized a model that offers insights into the subjective health experience (SHE) of patients and their associated needs [15]. These needs define the appropriate healthcare interventions that can be utilized alongside conventional medical treatments in the context of tailored care. Bloem and Stalpers define SHE as: “An individual’s experience of physical and mental functioning while living his life the way he wants to, within the actual constraints and limitations of individual existence.” [15, p. 8]. Two psychological determinants, namely acceptance and perceived control, are intrinsically associated with SHE. A heightened level of acceptance, reflecting the degree to which patients can integrate their health status into their daily life, and control, illustrating the extent to which patients perceive opportunities to improve their health condition, augments the positivity of health perception. Acceptance and control, both measured using three questions, form the foundation for a segmentation model that prescribes who necessitates specific interventions and when, establishing a framework to initiate tailored care (Fig 1).

**Fig 1:**
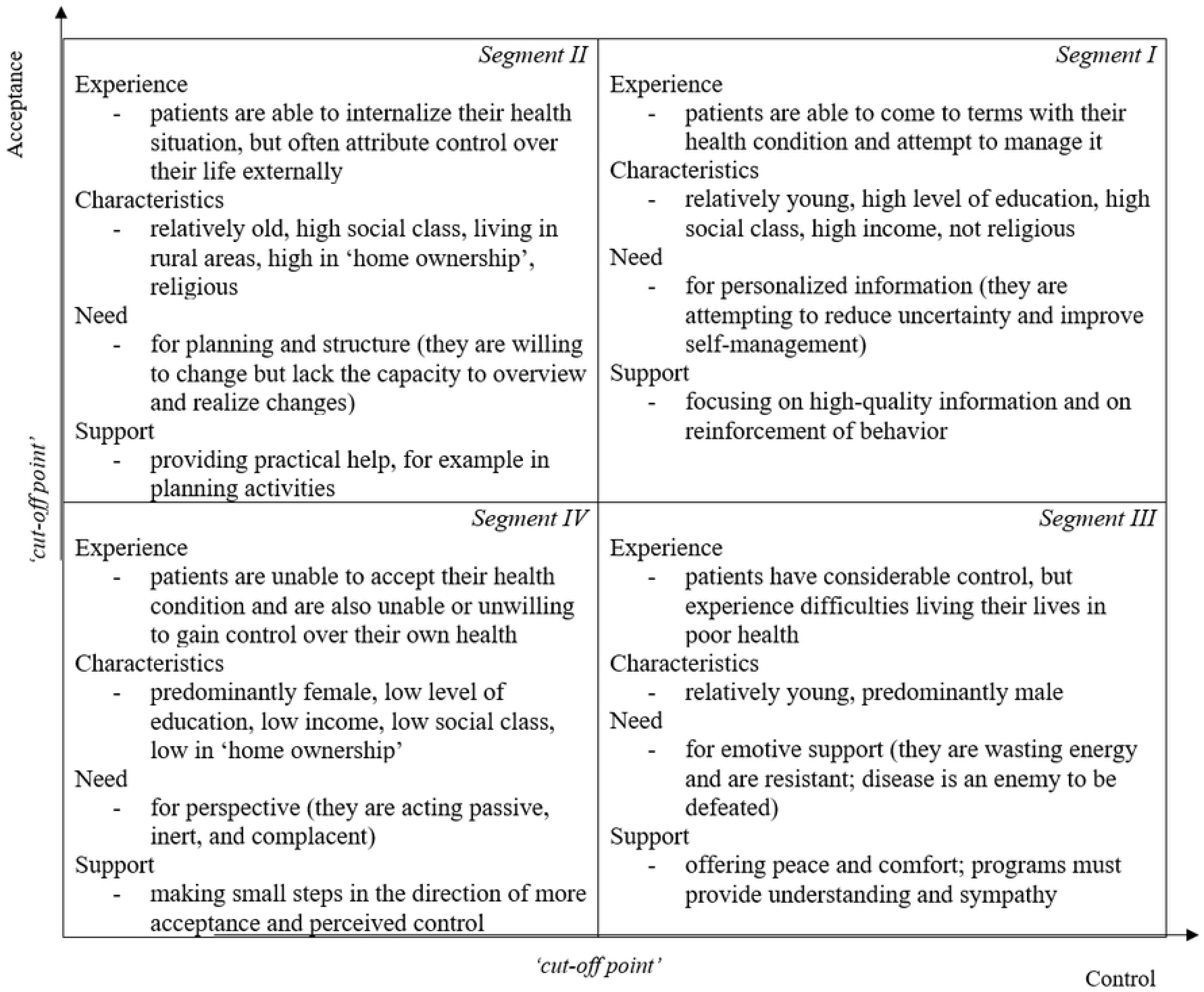
SHE-model based on Bloem, S. & Stalpers [15].

This SHE-model is inherently dynamic: health perceptions, acceptance, and perceived control can change over time due to a plethora of factors. Consequently, appropriate care modalities should exhibit adaptability over time.

Several studies have been conducted utilizing the SHE-model. For instance, the segments within this model have been further differentiated based on demographic and socio-economic variables [13], and in relation to vitality within older individuals [20]. In addition, health perceptions have been mapped across diverse disease domains [21], and the empirical relationship between health perception and health behaviour has been established [22]. Furthermore, in two distinct studies (cross-sectional and longitudinal), the model was validated for an IBD cohort [23,24].

While the SHE-model offers a guide for tailored care, it has not yet been adopted to cater to the distinctive needs of patients diagnosed with immunological disorders of various professional disciplines. We hypothesize that the needs for supportive care in this group of diseases will be comparable to each other as they have overlap in pathogenesis and treatment they encompass all lifelong chronic diseases with impact on social life and work capacity. Thus, this qualitative study aims to ascertain nuanced insights into the behaviour, questions (challenges), and specific needs of patients with six common immunological conditions guided by the SHE-model, thereby refining the personalized supportive care framework. It is hypothesized that the empirical outcomes derived from this research study will provide guidance to healthcare professionals in hospital settings to further enhance and more efficiently integrate this care into daily clinical practice.

### Method

This study is part of an extensive research project aiming to offer and monitor more personalized care related to immunological diseases and treatments based on Patient Reported Outcome Measures (PROMs). Additionally, it sought to enhance collaboration among the departments of gastro-enterology, rheumatology, and dermatology. A proposal for this research was submitted to the local Medical Ethics Committee at the Medical Centre Leeuwarden in May 2020. The proposal was approved and classified as not subject to the Medical Research Involving Human Subjects Act (nonWMO in Dutch). The recruitment period for this study was between May 1^st^ 2020 and April 18^th^ 2022.

### Study design

Collaborative discussions were initiated with both healthcare professionals and patients from three different disciplines, namely gastro-enterology, rheumatology, and dermatology, from The Medical Centre Leeuwarden, a prominent regional hospital in The Netherlands.

Both the healthcare professionals and patients with an immunological condition characterized the behaviour, questions (challenges), and specific needs of patients from the different segments of the SHE-model. For the patient groups, discussions also delved deeper into their disease perceptions. With both the healthcare professionals and the patients, an inventory was drawn up of the types of support offered by the hospital treatment teams. These modalities were subsequently allocated to one of the study’s four predefined segments.

### Participants and procedure

A total of nineteen healthcare professionals across three specialized teams participated in the research: an RA/SpA team comprised of three physicians and five nurses; an IBD team consisting of one physician and five nurses; and a PsO/HS team made up of one physician and four nurses. Length of service and gender varied among the participants.

Three groups of patients diagnosed with an immunological condition were interviewed. The RA/SpA group consisted of four patients diagnosed with RA and two with SpA. The IBD group included three patients with Crohn’s disease and four with Ulcerative Colitis. Finally, the PsO/HS group encompassed five patients with PsO and two with HS. Overall, eighteen participants were interviewed, of which ten were female and eight were male. The duration of their conditions ranged from two to fifty years, with participant ages varying between twenty-two and seventy-four years.

Group discussions were convened with all three of the healthcare professional teams, the RA/SpA patient group and the IBD patient group. The PsO/HS patient group was engaged in individualized interviews via Microsoft TEAMS (a logistical shift necessitated by the residual constraints of the COVID-19 pandemic, which prohibited in-person hospital interactions at the time). The discussions with healthcare professionals were executed first.

All patient participants provided explicit written authorization, through the written signing of an ‘informed consent’ form, allowing for the anonymized utilization of their interview data to enhance care approaches and for scientific publication. The study objectives and methodology were elucidated in supplemental documentation accompanying the consent form.

Three physicians (each of all three disciplines) involved in the research undertook the recruitment process, verbally inviting healthcare professionals and extending email invitations to patient participants. Participation was voluntary, and no compensatory incentives were disbursed.

### Guided discussion protocols

The structured group discussions, both with healthcare professionals and patients diagnosed with an immunological disorder, adhered to a predefined checklist. The structure of these interviews and the resultant data outputs are outlined in table 1 and 2. Notably, the segment descriptions provided by healthcare professionals served as input for the discussions with the individual participants. All interview sessions started with a preliminary introduction. This entailed an explanation of the research objectives and a reassurance regarding participant privacy and anonymity. All patient participants then provided formal consent to partake, as evidenced by the signing of an ‘informed consent’ document. Each session concluded with an overview of the subsequent steps in the research process.

**Table 1:** Structure of interview and output for healthcare professionals.

|  |  |
| --- | --- |
| Short explanation of the SHE-model<br>Describe each segment individually in terms of behaviours, questions, and needs. Constantly refer to your own professional experience, envisioning individuals who typically fit into each segment. Segments were defined either in small groups or individually, and the results were subsequently shared with the larger group.<br>Inventory of provided services<br>What array of services do you offer to support and guide patients? Information was collected collaboratively and documented on a flip chart. | Output<br>Description of the segments. A framework for differentiating healthcare modalities.<br>An inventory of available services was compiled. |
| Assigning support to segments (previous descriptions were visible)<br>What type of support is typically suited for which segment? A group discussion was conducted to determine this. | Consensus was achieved within the group based on discussion regarding the allocation of support across various segments. |

**Table 2:** Structure of interview and output for patients with immunological disorders.

|  |  |
| --- | --- |
| Top-of-Mind characteristics:<br>What comes to mind when you think about your [condition] and health? Outcome was documented on flip charts.<br>Select the three most significant words for you and describe what they mean in your daily life. This was an individual task, followed by a group discussion.<br>Health Ladder (Visual ladder with 11 Steps, brief explanation):<br>Describe an individual at the bottom and top of their health ladder. What does the person need to ascend the ladder? What does the hospital provide? This was discussed collectively.<br>Explanation of segmentation model (short video) + Brief description:<br>For each segment (visual provided with a brief description based on input from healthcare professionals), what services does the MCL offer for this individual? This was an individual task, followed by group discussion.<br>Note: In TEAM interviews, words were typed and displayed on the screen; the video was not shown. | Output:<br>Reflection on what the condition meant for the individual in their daily life; the aspects that played a role in this.<br>Insights into behaviour, questions, and needs; what was required; what was offered. (Note: High on the ladder corresponds with segment 1; low on the ladder corresponds with segment 4)<br>Insights into forms of support per segment. |

### Analytical approach

Interview transcripts were systematically analysed using the matrix method as proposed by Groenland to identify meaningful patterns [25]. Initially, a matrix was devised, designating rows for healthcare professionals and patients diagnosed with immunological disorders, while columns represented specific interview questions. Direct participant responses were transcribed and populated within this matrix.

In a subsequent phase, a thematic matrix was established to categorize and consolidate themes across various interview questions. Through quantitative tallying, predominant themes were differentiated, simultaneously elucidating the variances and commonalities both within and between participant groups.

This comprehensive analysis of the behaviours, questions, and needs specific to patients with immunological disorders, guided how tailored care strategies (provided by the hospital) could be distributed across the segments (also illustrated by Averill [26].

## Results

### Behavioural patterns, questions, and needs

The four segments of the SHE-model were subdivided into the categories: Cognition and behaviour, questions and dilemmas, and specific needs, based on the findings of the structured interviews with both the healthcare professional teams and individuals with an immunological condition (RA/SpA, IBD, or PsO/HS; outlined in Table 3). The results showed a considerable overlap both within and between participant groups and were therefore combined into one table. Relationships between the three characteristics, signifying a higher level of mutual relevance, are depicted in the table using dashed lines.

**Table 3:**
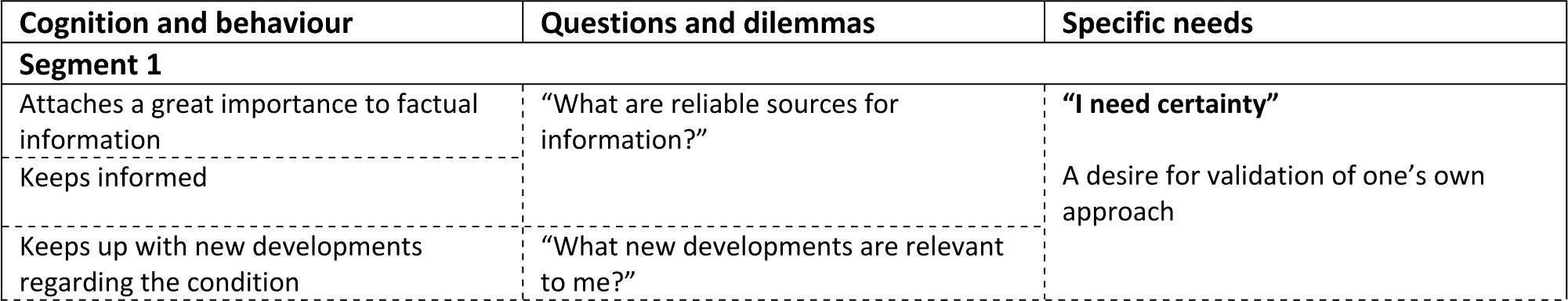
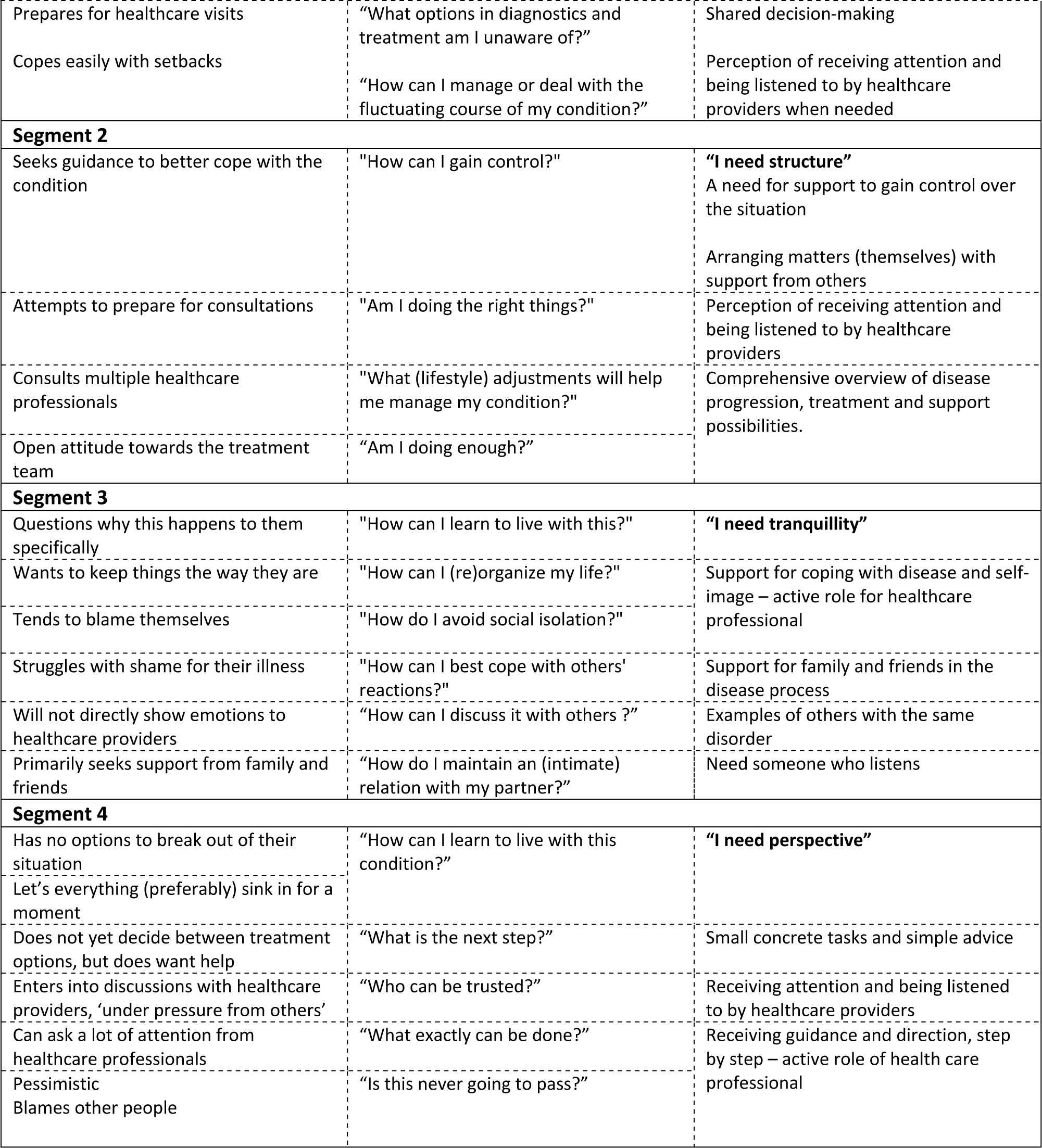
SHE-segment characteristics based on discussions with both healthcare professionals and patients (RA/SpA, IBD, PsO/HS)

| Cognition and behaviour | Questions and dilemmas | Specific needs |
| --- | --- | --- |
| **Segment 1** |  |  |
| Attaches a great importance to factual information<br>Keeps informed | "What are reliable sources for information?" | **"I need certainty"**<br>A desire for validation of one's own approach |
| Keeps up with new developments regarding the condition | "What new developments are relevant to me?" |  |
| Prepares for healthcare visits | "What options in diagnostics and treatment am I unaware of?" | Shared decision-making |
| Copes easily with setbacks | "How can I manage or deal with the fluctuating course of my condition?" | Perception of receiving attention and being listened to by healthcare providers when needed |
| **Segment 2** |  |  |
| Seeks guidance to better cope with the condition | "How can I gain control?" | **"I need structure"**<br>A need for support to gain control over the situation<br>Arranging matters (themselves) with support from others |
| Attempts to prepare for consultations | "Am I doing the right things?" | Perception of receiving attention and being listened to by healthcare providers |
| Consults multiple healthcare professionals | "What (lifestyle) adjustments will help me manage my condition?" | Comprehensive overview of disease progression, treatment and support possibilities. |
| Open attitude towards the treatment team | "Am I doing enough?" |  |
| **Segment 3** |  |  |
| Questions why this happens to them specifically | "How can I learn to live with this?" | **"I need tranquility"** |
| Wants to keep things the way they are | "How can I (re)organize my life?" | Support for coping with disease and self-image – active role for healthcare professional |
| Tends to blame themselves | "How do I avoid social isolation?" |  |
| Struggles with shame for their illness | "How can I best cope with others' reactions?" | Support for family and friends in the disease process |
| Will not directly show emotions to healthcare providers | "How can I discuss it with others?" | Examples of others with the same disorder |
| Primarily seeks support from family and friends | "How do I maintain an (intimate) relation with my partner?" | Need someone who listens |
| **Segment 4** |  |  |
| Has no options to break out of their situation | "How can I learn to live with this condition?" | **"I need perspective"** |
| Let's everything (preferably) sink in for a moment |  |  |
| Does not yet decide between treatment options, but does want help | "What is the next step?" | Small concrete tasks and simple advice |
| Enters into discussions with healthcare providers, 'under pressure from others' | "Who can be trusted?" | Receiving attention and being listened to by healthcare providers |
| Can ask a lot of attention from healthcare professionals | "What exactly can be done?" | Receiving guidance and direction, step by step – active role of health care professional |
| Pessimistic<br>Blames other people | "Is this never going to pass?" |  |

Legend: Dashed lines are used to indicate relationships between the three characteristics.

In segment 1, the key cognition and behaviour characteristic is their coping with the consequences of their chronic inflammatory disease. These patients display a high degree of openness when discussing their conditions with others (healthcare professionals, loved ones) and deal with comments regarding their disease in a proper and sensible manner. Overall, they attempt, whether with success or difficulty, to integrate the condition into their lives and engage in activities that align with their capabilities. An intrinsic motivation to remain well-informed is evident in their proactive acquisition of (novel) information pertaining to their conditions. They pose (critical) questions to both themselves and healthcare professionals. Furthermore, they actively obtain information from online sources to bring to their consultations. They wish to be actively involved regarding decisions about treatment and support. This group identifies with the concepts of ownership and self-management, and they seek external validation from healthcare professionals and loved ones to affirm their adaptive approach to managing their conditions.

In the second segment, patients exhibit a positive attitude towards their condition, aiming to optimize their well-being actively, yet continually searching for guidance to deal with the disease in a better way. Similarly to those in segment 1, these patients try to integrate their conditions and potential symptoms into their lives as well as possible. However, they frequently lack an overview of possible interventions. This group more frequently questions whether they are doing the right things to improve their quality of life. They are very willing to seek and accept assistance from others. Providing structured support and guidance are paramount in aiding these patients to manage their conditions effectively.

The third segment involves patients who cannot accept their disease and are not able to adapt to their condition. This segment predominantly comprising patients who have recently been diagnosed or whose circumstances have recently deteriorated. These respondents display a tendency to seek support from involved loved ones, although often suffering with feelings of shame. These patients harbour questions primarily concerning the management of their altered situation, both in relation to themselves and their environment. Immediate assistance in coping with the condition and self-perception, with the aim of getting into balance, is crucial for them.

In the fourth segment, was depicted by the respondents as patients who have relinquished hope. They have no perspective of how they can cope with the perceived impact of their disease and improve their QoL. Pervasive pain, fatigue, and other symptoms are significant impediments in their lives. They tend to withdraw from their social environment, driven by feelings of helplessness and the burden of shame. Managing comments, whether well-intentioned or not, becomes a challenge for them. This withdrawal results in more frequent feelings of isolation, misunderstanding, and melancholy. Their predominantly inert disposition poses challenges in motivating them to take proactive measures. Central to their concerns is the question of ‘what’s next?’. This question is particularly significant when they are faced with numerous lifestyle guidelines to adhere to. In light of these challenges, incremental progress through small tasks and guidance may lead to a renewed perspective. Engaging in activities, witnessing and experiencing results, and thus potentially regaining a sense of purpose are key to moving forward for patients from this segment.

Overall, the segment characteristics across the six immunological disorders displayed considerable overlap. Despite the apparent distinctions between these conditions, notable similarities exist in patients’ challenges, resulting in similar behaviours, questions, and needs. For instance, all patients consistently mentioned experiencing pain, while fatigue was reported by both RA/SpA and IBD patients. Pain and fatigue greatly impact daily activities, including home responsibilities, work, and social interactions. Furthermore, patients from all disorders struggle with feelings of shame, though its manifestation varies. For instance, reluctance to ask other people for help (e.g., RA/SpA), embarrassment due to frequent and unexpected need for visiting the toilet and confronted by faecal incontinence (e.g., IBD); shame stemming from feeling filthy and feeling perceived as such by others (e.g., PsO/HS).

### Tailored care

In Table 4, a comprehensive overview is presented, delineating the various forms of hospital-provided support for each segment of the SHE-model, as identified and categorized by healthcare professionals and patients (RA/SpA, IBD, or PsO/HS). It should be noted that the types of support discussed in the interviews were explicitly (and logically) related to the specific immunological disorders, but were generalized in the overview (except in cases where the form of support was uniquely applicable to a particular condition). Furthermore, during the discourse, at the initiation of the moderator, additional criteria were elucidated to refine the categorization of supportive care modalities. The study participants delineated between the ‘WHAT’, referring to the specific needs of patients diagnosed with immunological disorders, and the ‘HOW’, the methodological approach of providing the supportive care. This led to the conceptualization of a structured framework that categorizes the types of supportive care offered by healthcare professionals (as highlighted in the bold text in Table 4), alongside the optimal methods tailored to each SHE-segment (as indicated in the non-bolded text in the middle column, and validated by checkmarks in the rightmost column of Table 4).

**Table 4:**
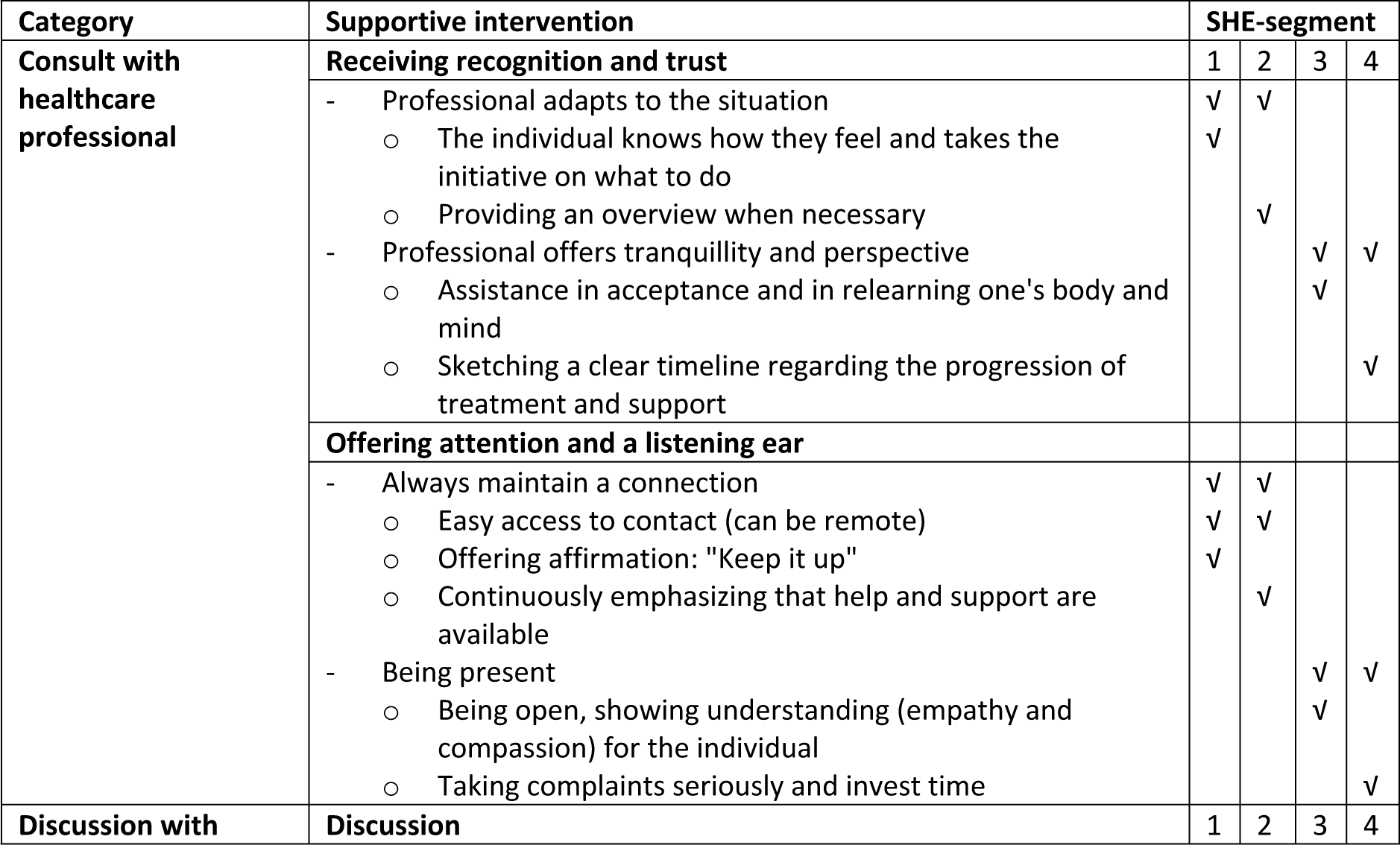
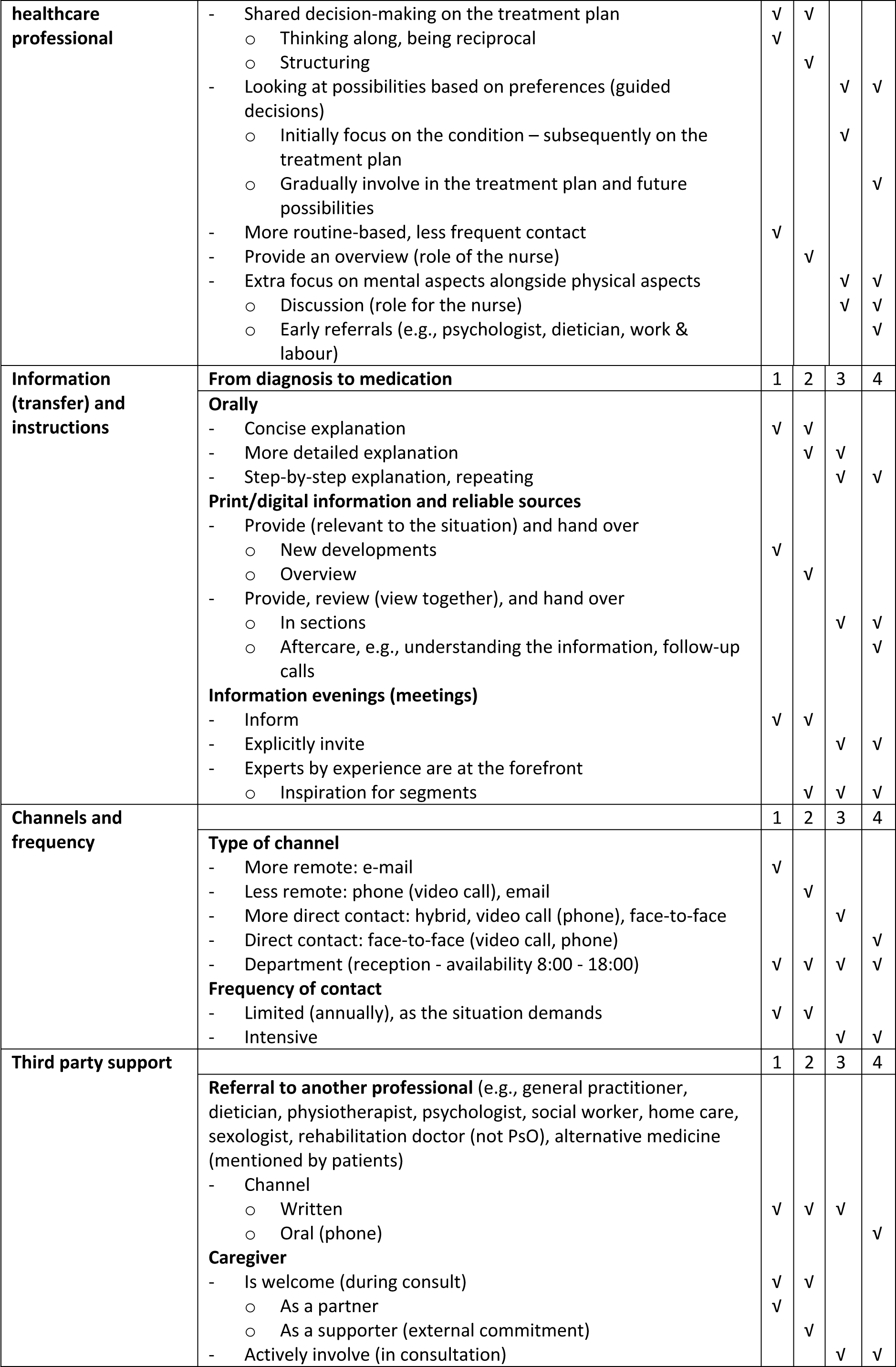
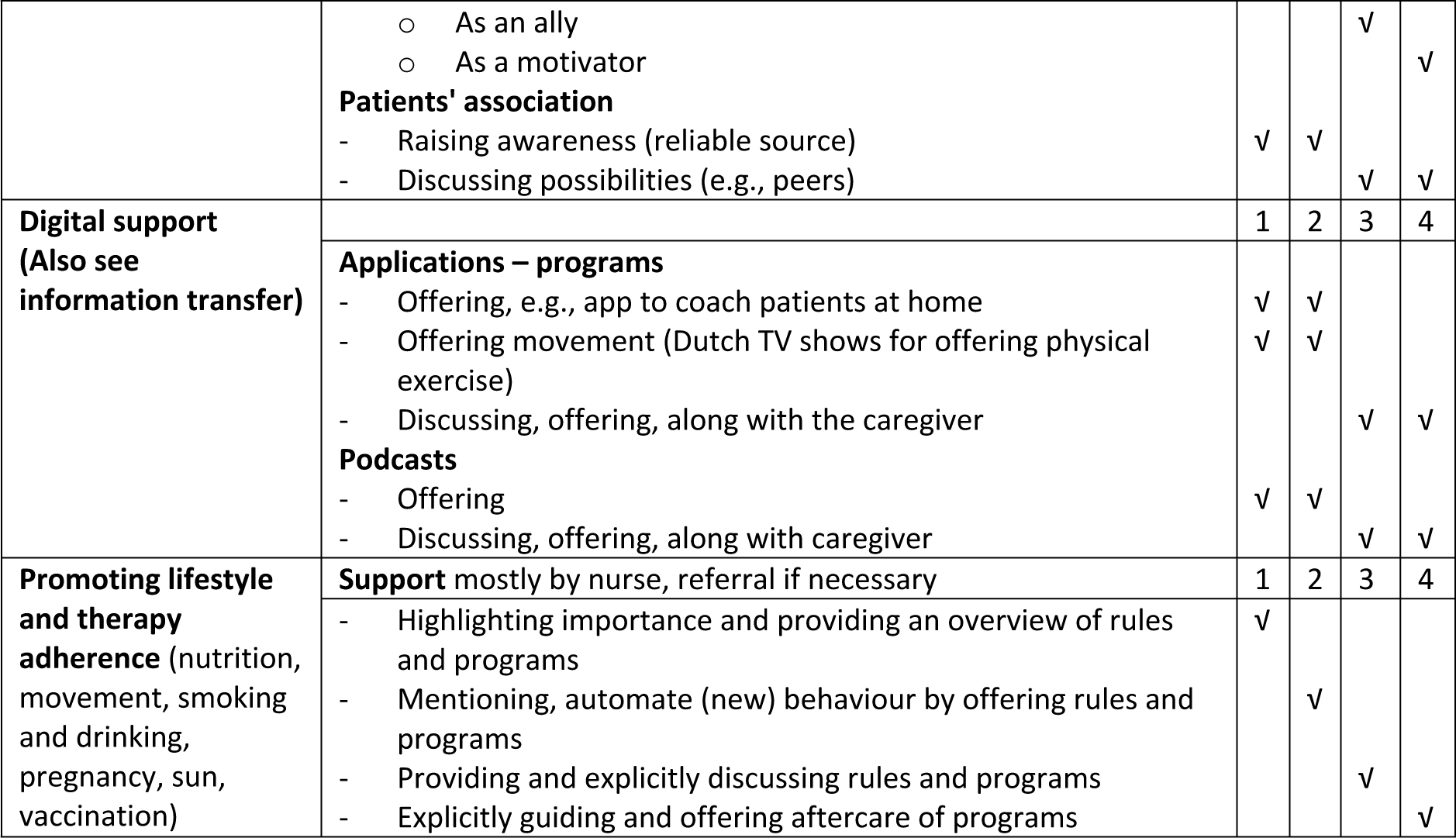
Tailored care framework based on discussions with both healthcare professionals and patients (RA/SpA, IBD, PsO/HS).

Legend: **bold** – mainly focused on ‘WHAT’ is offered, not bold – mainly focused on ‘HOW’ it is offered; numbers in right column refer to segments.

The participants, comprising both healthcare professionals and individual patients, occasionally found it challenging or non-intuitive to allocate specified forms of support to one or multiple segments. In initial assessments, respondents from both cohorts frequently indicated that universally relevant information is required for all segments, however, associations between the manner of information provision and specific patient segments were spontaneously highlighted by some respondents. For instance, offering information upon an individual’s explicit request was generally attributed to patients in segment 1, tailoring and structuring of information were deemed more suitable for patients falling under segment 2 and a more intensive engagement in discussing the available information was perceived as particularly beneficial for patients categorized within segments 3 and 4.

Furthermore, participants proposed that the distribution of information could either be furnished for the individual to review independently at home, or be interactively discussed with the individual during a clinical consultation. Additional variations included the potential segmentation of the information into discrete structured components, allocating more or less time during consultations for discussing this information, and the selection of appropriate communication channels both prior to and during the consultation. These nuanced insights provided respondents with further guidance on how to tailor supportive care interventions across different patient segments.

During the discussions, particularly among healthcare professionals, it became evident that the degree of an individual’s acceptance of their health condition fundamentally influences the preferred strategy for supportive care (the ‘HOW’). Patients who are further along in accepting their medical condition (segment 1 and 2) are more responsive to a reactive care approach, as opposed to those with a lower level of acceptance (segment 3 and 4), who notably benefit from a proactive model of care.

The high-acceptance cohort demonstrated a tendency toward self-initiative and greater autonomy in managing their care needs. They often independently coordinate their healthcare and engage in a more reciprocal relationship with healthcare providers. For these patients, shared decision-making emerges as a logical and effective collaborative care strategy. In contrast, the low-acceptance group, exhibits a lack of initiative, necessitating external guidance for organizing appropriate care and with decision-making. It is imperative to gain insights into their unique care preferences. Utilizing these insights, healthcare professionals are better equipped to offer a curated set of options, facilitating a process we call ‘guided decision-making’.

In addition, during discussions with healthcare professionals, it emerged that patients who are at a lower level of acceptance with regard to their health conditions (segments 3 and 4) experience greater difficulty in integrating their disorder into their life. Consequently, participants emphasized that prior discussing treatment modalities, considerable focus should be allocated to exploring the inherent implications of the disorder for the individual in question. The process surrounding grief, acceptance, and active involvement is universally experienced by patients, irrespective of their specific condition. Yet, this pivotal process often remains inadequately assessed by healthcare professionals, who tend to prioritize the quantifiable aspects of disease activity during consultations. In the process of identifying suitable care for each segment, patients with immunological disorders particularly focused on the nature of ‘contact’ with healthcare professionals (i.e., WHAT is needed?). The unanimous consensus among the respondents was that ‘recognition’ serves as an indispensable element. Recognition fosters the establishment of trust, creating the groundwork for a sustainable, long-term professional relationship. There exists a perpetual need to rely on professionals for varying forms of assistance and support—ranging from treatment and medication to discussions addressing physical and psychological issues. Over 50% of the respondents of the patients groups accentuated that this personalized recognition, which honours their intrinsic identity, facilitates mutual comprehension, thereby enhancing the overall quality of the healthcare they receive. Several illustrative examples were cited by the respondents, stemming from their personal experiences and unique medical conditions. These examples included trust-based consultations with gastroenterologists concerning pregnancy while using medication, re-evaluations of treatment protocols with rheumatologists due to increasing fatigue and diminished occupational performance, and candid discussions with dermatologists about the emotional toll of visible skin lesions, especially during summer months. Across all these scenarios, the principles of ‘recognition’ and ‘trust’ in healthcare professionals emerged as pivotal factors enabling efficacious healthcare provision.

A number of patients emphasized their perception of ‘vulnerability’, attributable to their ‘limited functional capabilities’ and the consequent ‘reliance on external assistance’. Moreover, participants identified an additional layer of ‘vulnerability’ arising from their (perceived) inferior status relative to healthcare providers. This perception was reported to be particularly prominent during engagements with physicians as compared to nurses, thereby impeding patients in talking completely openly during consultations. An essential element for attaining recognition, as underscored predominantly by patients with immunological disorders, centres on the healthcare professional’s capacity for ‘non-judgmental’ listening. Such an approach fosters the essential level of patient-centric attention and establishes listening as a ‘foundational precondition for achieving empathic understanding’.

These dialogues and insights contributed to a nuanced differentiation of healthcare service delivery strategies (HOW to implement this) across the segments (Table 4).

In the context of support facilitated by third parties, healthcare professionals exclusively differentiated among various communication channels employed during the referral process. Specifically, written referrals were predominant in patient segments 1 through 3, while telephonic consultations were reserved for segment 4, as outlined in Table 2. Moreover, healthcare professionals delineated nuanced roles for caregivers of patients afflicted with immunological conditions. In segment 1, caregivers act as ‘equal partners,’ providing affirmation and encouragement, whereas in segment 2 they serve as ‘supporters,’ helping patients adhere to appointments and acting as external commitments. In segment 3, caregivers play the role of ‘empathizers’ who understand and engage in discussions, and in segment 4, they function as ‘motivators’ that facilitate incremental progress. During the consultations, caregivers should be accordingly educated and guided on this aspect.

Lastly, healthcare professionals in particular, noted the limited availability of digital support tools (data collected before COVID-19 pandemic), suggesting a need for their development, possibly on a hospital-wide scale.

## Discussion

Drawing upon empirical insights into the behavioural patterns, questions, and specific needs of patients afflicted with immunological disorders, segments within the SHE-model were allocated for this cohort (Table 3). Therapeutic interventions from multidisciplinary healthcare teams were allocated across these segments, focusing on both ‘WHAT’ is offered and ‘HOW’ it is delivered (Table 4).

Patients notably prioritize ‘receiving attention’ and ‘active listening’ from medical professionals. These aspects are intrinsically linked to patient recognition, a concept that encompasses cognitive, emotional, and behavioural dimensions. In scientific literature, attention and active listening are frequently cited as pivotal indicators of the quality of the interaction between healthcare professionals and end-users [27–29]. For instance, it has been evidenced that attention and active listening contribute to a multitude of favourable outcomes, including improved patient guidance, heightened patient motivation, reduced frequency of healthcare service utilization, and elevated levels of patient satisfaction [30–32]. While the importance of ‘attention’ is commonly emphasized, the dimension of ‘listening’, as identified by study participants, is often comparatively underemphasized. This includes aspects such as unbiased listening and refraining from proposing immediate solutions. Rogers allocated specific focus on these dimensions in his therapeutic interactions and formulated targeted methodologies to address them [33]. Van de Pol operationalized this concept through the development of a ‘listening thermometer’, a pragmatic instrument designed to facilitate the transition from the act of listening to the state of acknowledgment [34]. This tool has demonstrated potential for straightforward applicability within a clinical setting.

Participants, both healthcare professionals and patients, frequently encountered challenges when attempting to categorize distinct forms of support across the segments. However, the dialogues during the study generated innovative approaches to differentiation, such as contrasting the mechanisms of ‘providing’ and ‘discussing’ information with patients. This offered participants new insights, that could be utilized as tools for more precise allocation of forms of support. Future research could benefit from a comprehensive list of such distinguishing mechanisms, which could subsequently be presented to participants when tasked with allocating different forms of support to the segments. Interestingly, the utilization of digital forms of support was seldom mentioned by the participants of all groups. This may be attributed to limited availability (this study was conducted partially pre-COVID-19) and lack of awareness among participants. As digital technology becomes increasingly important within the framework of tailored care, it will be crucial for innovation departments to focus on this area.

All study participants exhibited a comprehensive understanding of the segmentation model and demonstrated its applicability in practice. This model will offer healthcare professionals directional insights for discussing various forms of support with patients. These discussions could range from ‘guided decisions’, where options align with the patients pre-established set of preferences, to ‘shared decision-making’, in which end-users exhibit greater initiative. Notably, the model is designed to be flexible rather than prescriptive; it accommodates the possibility for both healthcare providers and end-users to override its segmentation criteria when deemed necessary. This ensures that adherence to algorithmic recommendations does not become overly rigid, thus maintaining the clinical judgment and individualized care central to effective healthcare provision.

### Strengths and Limitations

One of the key strengths of this study is its ecological validity, which lies in the involvement of both healthcare professionals (physicians and nurses) as well as patients suffering from six distinct immunological disorders. This study provides a framework, based on behaviour, questions, and healthcare needs, offering a valuable resource for both inspiration and evaluative metrics. Conversely, the study exhibits some limitations. Primarily, the evaluation is confined to the healthcare services currently offered by three treatment teams in one peripheral healthcare institution. While this was the study’s original aim, the findings may not universally extend to other medical teams or healthcare settings. Thus, additional or innovative initiatives not examined here may potentially better meet the healthcare needs of the patient population studied. Moreover, the study does not explore the qualitative aspects of the different forms of patient support offered. While this was not the research focus, the quality of such support would be a determinant of the overall quality of patient care and guidance.

Furthermore, the aforementioned ‘vulnerability’— stemming from patients’ perceived inferiority to healthcare providers—may not only inhibit open dialogue during consultations but could also influence their responses during this study’s interviews.

### Practical implications

The further operationalization of the SHE-model demonstrates that forms of support can be differentiated into appropriate care in a relatively straightforward manner. By integrating this model into the hospital’s Electronic Health Records (EHR), patients can receive more tailored guidance based on their levels of acceptance and control. The process can be monitored in real-time. Since subjective experiences and their determinants can vary, it is essential to measure these at regular intervals. The EHR should include a list of available support forms and a feature to record who receives what type of support. Over time, evaluations can be conducted based on this real-world data. Importantly, clear dashboards should be developed for both healthcare professionals and patients. In this manner, care can be optimized based on data, thereby enhancing the patient’s perception of health and overall quality of life. This serves as the foundation for efficient, appropriate care.

Further development and implementation of digital tools are essential. A recent study has also proven the validity, applicability and value of the SHE-model in the clinical care environment through manual procedures, but concluded that to achieve consistency and maximize effect, digital tools are needed [35]. Innovation departments within the hospital can play a pivotal role in this, possibly across various medical specialties.

This study has been executed in a clinical care/practice environment, but the suggestions for and the value of tailored care are also valid for the clinical studies in clinical research. A clinical study may be less complex (monopharmacy versus polypharmacy) and more controlled due to the strict study protocol, but in essence it still is about a healthcare professional and a patient working together to bring the treatment of a condition to a successful closure. On average and across all conditions clinical studies tend to suffer from a steady 25% early drop-out rate, which is an extreme form of non-adherence. A recent review shows that almost 70% of the protocol deviations can be linked to non-adherent behaviour of the patient [36]. In clinical studies early drop-out and protocol deviations lead to extended timelines, higher costs, lower efficacy, and missing clinical datasets. Tailored support based on the SHE model of patients during clinical studies may help to make patient adherence better and these studies more efficient and cost effective.

### Future Research

Future research should be targeted toward exploring a broader range of clinical domains. This will provide an opportunity to delineate any disease-specific differences or trends across varied healthcare contexts. Furthermore, focus should be on understanding the dynamic nature of disease processes (progress of conditions and their treatment), examining how patient behaviour and needs evolve over time and what implications this has for healthcare provision. Additionally, gaining insights into the effectiveness of healthcare interventions is vital. This will involve determining which approaches are most effective, which are less so, and what modifications could lead to better outcomes.

Furthermore, patients with new onset chronic immunological diseases all undergo processes as sorrow, understanding, acceptation, and lifestyle changes. Ultimately, using a general approach the SHE-model could facilitate adequate supportive treatment for patients grouped on segmentation, irrespective of their type of immunologic disease.

### Conclusion

Currently, detection of specific needs or type of approach for patient specific support is largely pending on personal sensitivity of the health care professional. This study has used the SHE-model to recognize and understand the personal need for supportive care for people with immunological conditions. Interestingly, this was general applicable to the six different diseases.

The findings led to the differentiation of multiple forms of supportive care across the SHE-segments, thus providing healthcare professionals evidence-based guidelines to tailor individualized treatment approaches. The core elements of effective supportive care identified in this study are attention, acknowledgment, and active listening, which are important factors in the provision and management of patient-centric, appropriate care.

## Data Availability

All relevant data are within the manuscript and its Supporting Information files.

## Acknowledgements

The authors would like to acknowledge the support of all staff at the participating facilities and also the participants who engaged in this project. Special thanks are extended to Damien S.E. Broekharst for his invaluable critical feedback that greatly enhanced the quality of this work.

## Abbreviations

EHR: Electronic Health Records
HS: Hidradenitis Supperativa
IBD: Inflammatory Bowel Disease
NonWMO: Not subject to the Medical Research Involving Human Subjects Act
PROMs: Patient Reported Outcome Measures
PsO: Psoriasis
QoL: Quality of Life
RA: Rheumatoid Arthritis
SHE: Subjective Health Experience
SpA: Spondylarthritis

